# Benefits and Cost of a $35 Insulin Cost-Sharing Cap for Medicare Patients

**DOI:** 10.1101/2022.01.06.22268864

**Authors:** Kao-Ping Chua, Joyce M. Lee, Joshua E. Tucker, Dominique Seo, Rena M. Conti

## Abstract

**BACKGROUND:** To improve insulin affordability, Congress is considering capping insulin cost-sharing to $35 per 30-day supply for Medicare patients. The potential benefits and cost of this cap are unclear. Additionally, it is unknown whether the benefits of this cap would vary between Medicare patients with type 1 versus type 2 diabetes.

**METHODS:** We conducted a cross-sectional analysis of the IQVIA Longitudinal Prescription Database, which reports prescriptions dispensed from 92% of U.S. pharmacies, and the Optum Clinformatics Data Mart, a national claims database from Medicare Advantage patients. The IQVIA analysis included patients who only had dispensed insulin prescriptions paid by Medicare in 2019. We estimated the proportion of Medicare patients who would benefit from an insulin cost-sharing cap of $35 per 30-day supply. Among these patients, we calculated the mean annual decrease in insulin out-of-pocket spending. We summed this decrease across patients to estimate the cap’s cost to the federal government. The Optum analysis included Medicare Advantage patients with diabetes and ≥1 dispensed insulin prescription in 2019. We used linear regression to compare the proportion of patients who would benefit from a $35 cap and annual savings among these patients by diabetes type, adjusting for demographic characteristics and payer type.

**RESULTS:** The IQVIA analysis included 2,227,229 patients who only had dispensed insulin prescriptions paid by Medicare in 2019. Mean (SD) age was 69.2 (11.4) years. The $35 cap would benefit 887,051 (39.0%) of patients, lowering annual insulin out-of-pocket spending by $338, from $687 to $349. Across all patients in the sample, aggregate savings (i.e., the cap’s cost to the federal government) would be $299,402,402, or a mean of $134.4 per patient. Among the 60,300 Medicare Advantage patients in the Optum Analysis, mean age was 72.6 (9.3) years; 2,686 (4.5%) had type 1 diabetes and 57,614 (95.6%) had type 2 diabetes. The $35 cap would benefit a higher proportion of patients with type 1 diabetes (64.0%) compared with patients with type 2 diabetes (59.4%). Among patients with type 1 diabetes who would benefit from the cap, annual savings would be greater ($284) compared with their counterparts with type 2 diabetes ($231; p<.001 in adjusted analyses for all comparisons).

**CONCLUSIONS:** A $35 insulin cost-sharing cap would benefit a sizable proportion of Medicare patients using insulin and may particularly lower out-of-pocket spending for patients with type 1 diabetes. The estimated cost of this cap to the federal government would be $134.4 per Medicare patient using insulin.

## INTRODUCTION

Improving insulin affordability is an urgent national priority.^1^ Insulin cost-sharing imposes financial barriers that may lead to underuse and poor glycemic control among patients with diabetes.^2^ To mitigate these barriers, Congress is currently debating whether to include a provision in the Build Back Better Act that would cap insulin cost-sharing to $35 per 30-day supply for all Medicare Part D beneficiaries.^3,4^ While a prior study estimated the benefits of a federal insulin cost-sharing cap for privately insured children and young adults with type 1 diabetes, the study did not estimate the cap’s cost, and generalizability of results to Medicare patients is uncertain.^5^

In this study, we used 2019 national prescription dispensing data to estimate the benefits and cost of a $35 insulin cost-sharing cap for Medicare patients. Additionally, using 2019 claims data from a national sample of Medicare Advantage patients, we determined whether the benefits of such a cap would differ among patients with type 1 versus type 2 diabetes.

## METHODS

### Data sources

In fall 2021, we conducted a cross-sectional analysis of two databases. The first was the 2019 IQVIA Longitudinal Prescription Database, which reports all prescriptions dispensed from 92% of U.S. retail pharmacies, 70% of mail-order pharmacies, and 70% of long-term care pharmacies. The database does not capture dispensing in closed pharmacies that only serve patients of a particular health care system, such as the Veterans Administration and Kaiser Permanente. For each insulin prescription, the database reports days supplied, the state in which prescribers practiced, out-of-pocket spending, and method of payment (cash, commercial, Medicare, and Medicaid). The database does not report patient race or ethnicity, diagnosis codes, or type of out-of-pocket payment (e.g., deductibles versus co-insurance). **Appendix 1** lists insulin products included in the IQVIA analysis.

Although IQVIA data are nationally representative, they lack the clinical information needed to differentiate between insulin use by patients with type 1 versus type 2 diabetes. Consequently, we also analysed the 2019 Optum Clinformatics Data Mart, which contains medical and pharmacy claims from Medicare Advantage enrollees across the U.S. Data include diagnosis codes and out-of-pocket spending for prescriptions via deductibles and co-payments. No patients in the database had co-insurance payments for prescription drugs. Because data were de-identified, the Institutional Review Board of the University of Michigan Medical School exempted this study from human subjects review.

### Study population

In the IQVIA analysis, we included patients for whom all insulin prescriptions dispensed in 2019 were written by prescribers practicing in one of the 50 U.S. states or the District of Columbia. We excluded patients with missing or invalid data for demographic characteristics, method of payment, days supplied, or out-of-pocket spending on any insulin claim in 2019. Finally, we limited to patients with insulin prescriptions only paid by Medicare. Patients with insulin prescriptions paid both by Medicare and other payers, including commercial insurance, Medicaid, and cash, were excluded.

In the Optum analysis, we included Medicare Advantage patients living in one of the 50 U.S. states or the District of Columbia who were continuously enrolled throughout 2018 and 2019, had ≥1 claim in 2018 containing an *International Classification of Disease, Tenth Revision, Clinical Modification* (ICD-10-CM) diagnosis code for type 1 diabetes (E10.xxx) or type 2 diabetes (E11.xxx), and had ≥1 insulin claim in 2019 (pharmacy claims with a national drug code for insulin). We excluded patients with missing or invalid data for demographic characteristics or days supplied. We required ≥1 diabetes diagnosis code in 2018 to limit to patients with established diabetes. We required ≥1 insulin claim in 2019 to limit to patients using insulin, thus excluding patients with type 2 diabetes who do not use this drug.

To classify whether patients had type 1 versus type 2 diabetes, we divided the number of times a diagnosis code for type 1 diabetes appeared on medical claims in 2018 by the number of times a diagnosis code for type 1 or type 2 diabetes appeared on these claims. If this proportion exceeded 0.5, we classified the patient as having type 1 diabetes; otherwise, we classified the patient as having type 2 diabetes. A prior validation study suggests that this definition has high positive predictive value for identifying patients with type 1 diabetes in administrative data.^6^

### Outcomes

In the IQVIA and Optum analyses, we calculated annual out-of-pocket spending across all insulin claims in 2019 for each patient. We then estimated this spending under a federal law that capped insulin cost-sharing to $35 per 30-day supply. When assessing the impacts of this cap, we made five assumptions. First, we assumed that the cap applied to all insulin prescriptions, regardless of insulin type or method of payment. Second, we assumed that the cap was applied on a per-prescription basis. Although a few existing caps collectively limit cost-sharing across all insulin prescriptions filled during a calendar month, including Colorado’s cap,^7^ per-prescription caps are more prevalent, perhaps owing to greater ease of implementation.^8^ Third, we assumed that the cap limited cost-sharing to the same level for prescriptions with days supplied between 1-30 days and that longer durations triggered a new cap. For example, maximum cost-sharing was $70 if days supplied in the prescription was 31-60 days. Fourth, we assumed the cap did not affect the number of insulin prescriptions filled by patients, an assumption supported by evidence that insulin use varies little with price.^9^ Finally, we assumed the cap did not affect if or when patients met deductibles or out-of-pocket maximums. This assumption was necessary because we lacked information on plan benefit design.

In the IQVIA analysis, we calculated the proportion of patients for whom the $35 cap would lower annual insulin out-of-pocket spending as well as mean annual savings among these patients. We summed savings across patients to estimate the cost of the cap to the federal government. In the Optum analysis, we used linear regression to assess whether diabetes type was associated with differences in the proportion of Medicare Advantage patients who would benefit from the cap and with differences in the magnitude of annual savings among patients who would benefit. Models controlled for age group, sex, and Census region of residence. Analyses used SAS 9.4 (SAS Institute), Stata/MP 14.2 (StataCorp), and two-sided hypothesis tests with α = 0.05.

## RESULTS

### Sample characteristics

In the IQVIA database, 6,495,240 patients met inclusion criteria, of which 574,501 (8.9%) were excluded owing to missing or invalid data for demographic characteristics, method of payment, days supplied, or out-of-pocket spending on any insulin claim in 2019 (see **Appendix 2** for details). Of the remaining 5,920,739 patients, 2,227,229 (37.7%) had insulin prescriptions only paid by Medicare. **Table 1** displays characteristics of these 2,227,229 patients. Mean (SD) age was 69.2 (11.4) years and 1,222,854 (53.7%) patients were female. Patients had a mean of 6.6 (5.7) insulin prescriptions and 305.2 (197.0) days supplied of insulin in 2019.

**Table 1.** Characteristics of Americans with dispensed insulin prescriptions only paid by Medicare in 2019, IQVIA Longitudinal Prescription Database

| **Characteristic** | **Number (%)** |
| --- | --- |
| **Number of patients** | 2,277,229 |
| **Age, mean (SD)** | 69.2 (11.4) |
| **Age group** |  |
| 17 and under | 6,769 (0.3) |
| 18 to 34 | 22,239 (1.0) |
| 35 to 54 | 192,835 (8.5) |
| 55 to 64 | 345,114 (15.2) |
| 65 and above | 1,710,272 (75.1) |
| **Gender** |  |
| Female | 1,222,854 (53.7) |
| Male | 1,053,931 (46.3) |
| Unknown | 444 (0.0) |
| **Region<sup>b</sup>** |  |
| Northeast | 387,766 (17.0) |
| Midwest | 513,013 (22.5) |
| South | 998,287 (43.8) |
| West | 378,163 (16.6) |
| **Days supplied of insulin, mean (SD)** | 305.2 (197.0) |
| **Annual number of insulin claims, mean (SD)** | 6.6 (5.2) |

<sup>a</sup>This refers to the region in which prescribers of the first insulin prescription during 2019 practiced. Our view of the IQVIA data did not report the state in which patients resided.

In the Optum database, there were 409,080 Medicare Advantage patients who lived in one of the 50 U.S. states or the District of Columbia, were continuously enrolled throughout 2018 and 2019, and had ≥1 claim for type 1 diabetes or type 2 diabetes in 2018. Of these patients, 60,303 (12.5%) had ≥1 claim for insulin in 2019. Three patients were excluded owing to missing or invalid data for demographic characteristics or days supplied, leaving 60,300 patients in the sample. Among the 60,300 Medicare Advantage patients, mean age was 72.6 (9.3) years; 34,028 (56.4%) were female, 31,395 (52.1%) resided in the South, 2,686 (4.5%) had type 1 diabetes, and 57,614 (95.6%) had type 2 diabetes. Mean annual number of dispensed insulin prescriptions was 8.0 (7.7). Mean days supplied of insulin was 342.8 (186.9) days.

### IQVIA analysis

A $35 cap would benefit 887,051 (39.0%) of Medicare patients using insulin, lowering their annual insulin out-of-pocket spending by $338 (from $687 to $349; **Table 2**). The estimated cost of the cap for patients in this sample was $299,402,402, corresponding to a mean of $134.4 per patient. For the 61.0% of Medicare patients who would not benefit from the cap, mean (SD) annual out-of-pocket spending would remain at $47 (89).

**Table 2.** Annual insulin out-of-pocket spending under a $35 federal cap for Medicare patients, 2019 IQVIA Longitudinal Prescription Database

| Category | Outcome | \$35 cap |
| --- | --- | --- |
| Patients who would benefit from cap | Number (% of patients with Medicare only) | 887,051 (39.0) |
| | Mean (SD) annual OOP spending without cap, \$ | 687 (606) |
| | Mean (SD) annual OOP spending with cap, \$ | 349 (224) |
| | Mean (SD) decrease in annual OOP spending with cap, \$<sup>e</sup> | 338 (473) |
| Patients who would not benefit from cap | Number (% of patients with Medicare only) | 1,290,178 (61.0) |
| | Mean (SD) annual OOP spending with or without cap, \$ | 47 (89) |

OOP – out-of-pocket.

### Optum analysis

In unadjusted analyses, the $35 cap would benefit a higher proportion of patients with type 1 diabetes (64.0%) than type 2 diabetes (59.4%; **Table 3**). Among patients with type 1 diabetes who would benefit from the cap, annual savings would be greater ($284) compared with their counterparts with type 2 diabetes ($231; p<.001 in adjusted analyses for all comparisons).

**Table 3.** Annual insulin out-of-pocket spending under a $35 federal cap among Medicare Advantage patients with type 1 versus type 2 diabetes, 2019 Optum Clinformatics Data Mart

| Population | Category | Outcome | \$35 cap |
| --- | --- | --- | --- |
| **Type 1 Diabetes**<br>**(n = 2,686)** | Patients who would benefit from cap | Number (% of patients with type 1 diabetes) | 1,719 (64.0%) |
| | | Mean annual OOP spending without cap, \$ | 640 (448) |
| | | Mean annual OOP spending with cap, \$ | 356 (235) |
| | | Decrease in mean annual OOP spending with cap, \$<sup>e</sup> | 284 (327) |
|  | Patients who would not benefit from cap | Number (% of patients with type 1 diabetes) | 967 (36%) |
| | | Mean annual OOP spending with or without cap, \$ | 135 (185) |
| **Type 2 Diabetes**<br>**(n = 57,614)** | Patients who would benefit from cap | Number (% of patients with type 2 diabetes) | 34,226 (59.4%) |
| | | Mean annual OOP spending without cap, \$ | 524 (393) |
| | | Mean annual OOP spending with cap, \$ | 292 (225) |
| | | Decrease in mean annual OOP spending with cap, \$<sup>e</sup> | 231 (269) |
|  | Patients who would not benefit from cap | Number (% of patients with type 2 diabetes) | 23,388 (40.6%) |
| | | Mean annual OOP spending with or without cap, \$ | 97 (150) |

OOP – out-of-pocket.

### Additional analyses

For interested readers, we estimated the benefits and cost of a $25 and $50 cap for Medicare patients. Additionally, we estimated the benefit of a $25, $35, and $50 cap for patients who only had dispensed insulin prescriptions paid by commercial insurance in 2019. The results of these additional analyses are included in **Appendix 3**.

## DISCUSSION

Using 2019 national pharmacy dispensing data, we estimate that a federal law capping insulin cost-sharing for Medicare patients to $35 per 30-day supply would benefit 39.0% of Medicare patients using this drug. For these patients, annual insulin out-of-pocket spending would decrease by $338, from $687 to $349. The cost of the cap to the federal government would be $299.4 million, or a mean of $134.4 per patient. Using claims data from a national sample of Medicare Advantage patients with diabetes, we found that a $35 cap would benefit a higher proportion of patients with type 1 diabetes compared with those with type 2 diabetes. Additionally, the magnitude of annual savings would be greater among the former.

Collectively, our findings suggest that a $35 insulin cost-sharing cap would benefit a sizable proportion of Medicare patients using insulin and may particularly lower out-of-pocket spending for patients with type 1 diabetes. Importantly, however, Medicare patients who would benefit from the cap would still pay an average of $349 per year for insulin, a level that may still be burdensome for many patients. Consequently, additional interventions may be needed to reduce cost-related barriers to insulin access. For example, because drug cost-sharing is inextricably tied to price, interventions to lower insulin prices might be considered, including price negotiation, a policy option currently under consideration by Congress.^1^

The primary strength of this study is its use of an all-payer, national prescription dispensing database. However, the study has limitations. First, we may have misclassified patients as having type 1 versus type 2 diabetes in the Optum analysis owing to our reliance on diagnosis codes. Second, it is unclear whether results from the Optum analysis generalize to all Medicare beneficiaries. Third, the cost of the cap to the federal government may have either been overestimated or underestimated. On the one hand, we analyzed data from 2019, the year before many Medicare Part D plans implemented a similar $35 insulin cost-sharing cap,^8,10^ suggesting the cost could be overestimated. On the other hand, analyses did not capture all Medicare patients who use insulin. For example, a Kaiser Family Foundation analysis suggested that 3.1 million Medicare patients used insulin in 2017.^11^ In contrast, our sample only included 2.2 million Medicare patients who used insulin in 2019, potentially because IQVIA data do not all include prescriptions, because we excluded patients who had insulin prescriptions paid both by Medicare and other payers, and because we excluded patients with missing data on insulin out-of-pocket spending. As a result of this limitation, the cost of the cap might be underestimated.

## CONCLUSIONS

Despite these limitations, our findings can inform ongoing debates regarding the benefits and cost of the insulin cost-sharing cap currently under consideration in the Build Back Better Act. If Congress implements a cost-sharing cap for Medicare patients with a different design than the one modeled in this study, we will update analyses to reflect the design of the cap.

## Data Availability

Data are unavailable because they are proprietary.

## Author Contributions

Dr. Chua had full access to all of the data in the study and takes responsibility for the integrity of the data and the accuracy of the data analysis.

*Study concept and design:* Chua, Lee, Tucker, Seo, Conti

*Acquisition of data:* Chua

*Analysis and interpretation of data:* Chua, Lee, Tucker, Seo, Conti

*Drafting of the manuscript:* Chua

*Critical revision of the manuscript:* Chua, Lee, Tucker, Seo, Conti

*Statistical analysis:* Chua, Tucker

*Study supervision:* Conti

