## Appendix for "Benefits and Cost of a $35 Insulin Cost-Sharing Cap for Medicare Patients"

**APPENDIX TABLE OF CONTENTS**

**Appendix 1.** List of insulin products included in the IQVIA analysis

**Appendix 2.** Sample inclusion and exclusion criteria in the IQVIA analysis

**Appendix 3.** Benefits and costs of a $25, $35, and $50 cap among Medicare and commercially insured patients

**Appendix 1.** List of insulin products included in the IQVIA analysis

These were products in Uniform Classification of Systems Level 3 (UCC3) 39100 or 39400.

| **GENERIC NAME** | **BRAND NAME** |
| --- | --- |
| INSULIN ASPART | INSULIN ASPART |
| INSULIN ASPART | INSULIN ASPART FLEXPEN |
| INSULIN ASPART | INSULIN ASPART PENFILL |
| INSULIN ASPART | NOVOLOG |
| INSULIN ASPART | NOVOLOG FLEXPEN |
| INSULIN ASPART | NOVOLOG PENFILL |
| INSULIN ASPART (WITH NIACINAMIDE) | FIASP |
| INSULIN ASPART (WITH NIACINAMIDE) | FIASP FLEXTOUCH |
| INSULIN ASPART (WITH NIACINAMIDE) | FIASP PENFILL |
| INSULIN ASPART PROTAMINE & ASPART (HUMAN) | INSULIN ASPART PROTAMINE/ |
| INSULIN ASPART PROTAMINE & ASPART (HUMAN) | NOVOLOG MIX 70/30 |
| INSULIN ASPART PROTAMINE & ASPART (HUMAN) | NOVOLOG MIX 70/30 PENFILL |
| INSULIN ASPART PROTAMINE & ASPART (HUMAN) | NOVOLOG MIX 70/30 PREFILL |
| INSULIN DEGLUDEC | TRESIBA |
| INSULIN DEGLUDEC | TRESIBA FLEXTOUCH |
| INSULIN DEGLUDEC-LIRAGLUTIDE | XULTOPHY 100/3.6 |
| INSULIN DETEMIR | LEVEMIR |
| INSULIN DETEMIR | LEVEMIR FLEXPEN |
| INSULIN DETEMIR | LEVEMIR FLEXTOUCH |
| INSULIN GLARGINE | BASAGLAR KWIKPEN |
| INSULIN GLARGINE | LANTUS |
| INSULIN GLARGINE | LANTUS FOR OPTICLIK |
| INSULIN GLARGINE | LANTUS SOLOSTAR |
| INSULIN GLARGINE | TOUJEO MAX SOLOSTAR |
| INSULIN GLARGINE | TOUJEO SOLOSTAR |
| INSULIN GLARGINE-LIXISENATIDE | SOLIQUA 100/33 |
| INSULIN GLULISINE | APIDRA |
| INSULIN GLULISINE | APIDRA SOLOSTAR |
| INSULIN ISOPHANE | ILETIN I NPH |
| INSULIN LISPRO | ADMELOG |
| INSULIN LISPRO | ADMELOG SOLOSTAR |
| INSULIN LISPRO | HUMALOG |
| INSULIN LISPRO | HUMALOG JUNIOR KWIKPEN |
| INSULIN LISPRO | HUMALOG KWIKPEN |
| INSULIN LISPRO | HUMALOG PEN |
| INSULIN LISPRO | INSULIN LISPRO |
| INSULIN LISPRO | INSULIN LISPRO KWIKPEN |
| INSULIN LISPRO PROTAMINE & LISPRO | HUMALOG MIX 50/50 |
| INSULIN LISPRO PROTAMINE & LISPRO | HUMALOG MIX 50/50 KWIKPEN |
| INSULIN LISPRO PROTAMINE & LISPRO | HUMALOG MIX 75/25 |
| INSULIN LISPRO PROTAMINE & LISPRO | HUMALOG MIX 75/25 KWIKPEN |
| INSULIN LISPRO PROTAMINE & LISPRO | HUMALOG MIX 75/25 PEN |
| INSULIN NPH (HUMAN) (ISOPHANE) | HUMULIN N |
| INSULIN NPH (HUMAN) (ISOPHANE) | HUMULIN N KWIKPEN |
| INSULIN NPH (HUMAN) (ISOPHANE) | HUMULIN N U-100 PEN |
| INSULIN NPH (HUMAN) (ISOPHANE) | NOVOLIN N |
| INSULIN NPH (HUMAN) (ISOPHANE) | NOVOLIN N FLEXPEN |
| INSULIN NPH (HUMAN) (ISOPHANE) | NOVOLIN N RELION |
| INSULIN NPH (HUMAN) (ISOPHANE) | NOVOLIN N U-100 |
| INSULIN NPH ISOPHANE & REG (HUMAN) | HUMULIN 70/30 |
| INSULIN NPH ISOPHANE & REG (HUMAN) | HUMULIN 70/30 KWIKPEN |
| INSULIN NPH ISOPHANE & REG (HUMAN) | HUMULIN 70/30 PEN |
| INSULIN NPH ISOPHANE & REG (HUMAN) | NOVOLIN 70-30 |
| INSULIN NPH ISOPHANE & REG (HUMAN) | NOVOLIN 70/30 |
| INSULIN NPH ISOPHANE & REG (HUMAN) | NOVOLIN 70/30 FLEXPEN |
| INSULIN NPH ISOPHANE & REG (HUMAN) | NOVOLIN 70/30 FLEXPEN REL |
| INSULIN NPH ISOPHANE & REG (HUMAN) | NOVOLIN 70/30 INNOLET |
| INSULIN NPH ISOPHANE & REG (HUMAN) | NOVOLIN 70/30 RELION |
| INSULIN NPH ISOPHANE & REG (HUMAN) | RELION 70/30 |
| INSULIN REGULAR (HUMAN) | AFREZZA |
| INSULIN REGULAR (HUMAN) | HUMULIN R |
| INSULIN REGULAR (HUMAN) | HUMULIN R U-500 (CONCENTR |
| INSULIN REGULAR (HUMAN) | HUMULIN R U-500 KWIKPEN |
| INSULIN REGULAR (HUMAN) | NOVOLIN R |
| INSULIN REGULAR (HUMAN) | NOVOLIN R FLEXPEN |
| INSULIN REGULAR (HUMAN) | NOVOLIN R RELION |
| INSULIN REGULAR (HUMAN) | RELION R |
| INSULIN REGULAR (PORK) | INSULIN STANDARD |
| INSULIN ZINC (HUMAN) | NOVOLIN L |
| None | HUMULIN N |
| None | HUMULIN R |
| None | NOVOLIN 70/30 |
| None | NOVOLIN N |
| None | NOVOLIN R |
| None | TRESIBA |
| None | ULTRALENTE ILETIN I |

**Appendix 2.** Sample inclusion and exclusion criteria in the IQVIA analysis

|  | **Number of patients remaining** | **Number of patients excluded** |
| --- | --- | --- |
| Number of patients who only had insulin prescriptions in 2019 from prescribers in one of the 50 states or the District of Columbia | 6,495,240 | N/A |
| Negative age | 6,487,579 | 7,661 |
| Invalid or missing days supplied for any insulin claim in 2019 | 6,487,554 | 25 |
| Unknown insurance type for any insulin claim in 2019 | 6,484,612 | 2,942 |
| Negative out-of-pocket spending for any insulin claim in 2019 | 6,484,610 | 2 |
| Missing out-of-pocket spending for any insulin claim in 2019 | 5,920,739 | 563,871 |
| Only had insulin prescriptions paid with Medicare | 2,227,229 | 4,268,001 |

**Appendix 3.** Benefits and costs of a $25, $35, and $50 cap among Medicare and commercially insured patients

“Commercial payment only” refers to patients who only had dispensed insulin prescriptions in 2019 that were paid via commercial insurance.

| **Population** | **Category** | **Outcome** | **$25 cap** | **$35 cap** | **$50 cap** |
| --- | --- | --- | --- | --- | --- |
| **Medicare payment only**  **(n = 2,277,229)** | Patients who would benefit from cap | Number (% of patients with Medicare only) | **970,575 (42.6)** | **887,051 (39.0)** | **602,442 (26.5)** |
|  |  | Mean (SD) annual OOP spending without cap, $ | **649 (594)** | **687 (606)** | **873 (642)** |
|  |  | Mean (SD) annual OOP spending with cap, $ | **257 (168)** | **349 (224)** | **516 (292)** |
|  |  | Mean (SD) decrease in annual OOP spending with cap, $^e^ | **392 (493)** | **338 (473)** | **357 (486)** |
|  | Patients who would not benefit from cap | Number (% of patients with Medicare only) | 1,306,654 (57.4) | 1,290,178 (61.0) | 1,674,787 (73.5) |
|  |  | Mean (SD) annual OOP spending with or without cap, $ | 34 (62) | 47 (89) | 89 (149) |
| **Commercial payment only**  **(n = 2,241,064)** | Patients who would benefit from cap | Number (% of patients with commercial only) | **1,110,672 (49.6)** | **815,839 (36.4)** | **568,788 (25.4)** |
|  |  | Mean (SD) annual OOP spending without cap, $ | **598 (874)** | **725 (984)** | **888 (1,129)** |
|  |  | Mean (SD) annual OOP spending with cap, $ | **215 (160)** | **284 (212)** | **365 (282)** |
|  |  | Mean (SD) decrease in annual OOP spending with cap, $^e^ | **383 (819)** | **441 (905)** | **523 (1,006)** |
|  | Patients who would not benefit from cap | Number (% of patients with commercial only) | 1,130,392 (50.4) | 1,425,225 (63.6) | 1,672,276 (74.6) |
|  |  | Mean annual OOP spending with or without cap, $ | 79 (109) | 114 (145) | 149 (184) |

Cost of caps to the federal government (for patients with Medicare payment only) or to commercial insurers (for patients with commercial payment only)

| **Population** | **$25 cap** | **$35 cap** | **$50 cap** |
| --- | --- | --- | --- |
| **Medicare payment only**  **(n = 2,277,229)** | 380,025,675 | 299,402,402 | 214,936,484 |
| **Commercial payment only**  **(n = 2,241,064)** | 425,782,470 | 359,566,773 | 297,426,510 |
